# Determinants of COVID-19 Vaccine Acceptability in Mozambique: The Role of Institutional Trust

**DOI:** 10.1101/2022.03.03.22271828

**Authors:** Bo Hu, Wei Yang, Paul Bouanchaud, Yolanda Chongo, Jennifer Wheeler, Sergio Chicumbe, Marcos Chissano

## Abstract

**Background:** Vaccination plays an imperative role in protecting public health and preventing avoidable mortality. Yet, the reasons for vaccine hesitancy are not well understood. This study investigates the factors associated with the acceptability of COVID-19 vaccine in Mozambique.

**Methods:** The data came from the three waves of the COVID-19 Knowledge, Attitudes and Practices (KAP) survey which followed a cohort of 1,371 adults in Mozambique over three months (N=3,809). Data collection was through a structured questionnaire using telephone interviewing (CAPI). Multilevel regression analysis was conducted to identify the trajectories of, and the factors associated with COVID-19 vaccine acceptability.

**Results:** There was great volatility in COVID-19 vaccine acceptability over time. Institutional trust was consistently and strongly correlated with different measures of vaccine acceptability. There was a greater decline in vaccine acceptability in people with lower institutional trust. The positive correlation between institutional trust and vaccine acceptability was stronger in younger than older adults. Vaccine acceptability also varied by gender and marital status.

**Conclusions:** Vaccine acceptability is sensitive to news and information circulated in the public domain. Institutional trust is a central driver of vaccine acceptability and contributes to the resilience of the health system. Our study highlights the importance of health communication and building a trustful relationship between the general public and public institutions in the context of a global pandemic.

## 1. INTRODUCTION

The speed with which vaccines have been developed, trialed, and approved during the global COVID-19 pandemic has demonstrated the global health community’s capacity to act quickly to contain the spread of SARS-CoV-2. Despite safe and effective vaccines becoming increasingly available (1,2) their acceptability and uptake in some populations have been lower than expected, with vaccine hesitancy recognized as a key barrier to uptake (3). Given the important role of vaccination in protecting public health, it is crucial to better understand the factors that may lead to vaccine acceptability or hesitancy. This is especially the case in the Global South, where evidence about COVID-19 vaccine acceptability in the population has to-date been limited.

Vaccine hesitancy has been identified by the WHO as one of the top 10 threats to world health (4) Multiple factors have been associated with increased vaccine hesitancy, including the proliferation of misinformation on social media (5), a lack of trust in public health institutions (in one French study)(6), mistrust in biomedicine (in Italy) (7), and sociodemographic differences (in the UK) (8). While the majority of research to date has been focused on the Global North, recent work in Mozambique suggests COVID-19 vaccine hesitancy was lower among health workers than others surveyed (86.6% and 64.9% saying they would take the vaccine, respectively) in one cross-sectional online survey (9). Evidence from other contexts has suggested that institutional trust is a key component of health service utilization. Building public trust in multisectoral actors through SBCC interventions therefore presents a potentially fruitful way to increase service utilization, including vaccine acceptability and uptake, which can lead to a more resilient health system.

According to the Mozambican national vaccination plan, the introduction of a safe and effective vaccine represents a national priority to alleviate the negative impact of the COVID-19 pandemic on the health and socioeconomic system. The aim of the national COVID-19 vaccination communication campaigns has been to disseminate prevention measures through traditional and new media including radio, television, newspapers, SMS, WhatsApp, and Facebook. The communication plan focuses on providing fast and up-to-date information on vaccination and explains, whenever possible, the process associated with the campaign, including vaccination sites, target groups, the safety and efficacy of vaccines, associated risks, and adverse reactions.

This paper uses three waves of longitudinal survey data to show how vaccine hesitancy has changed over six months in Mozambique, and what factors were associated with that change. We pay special attention to the influence of institutional trust. Institutional trust is a modifiable factor. A focused investigation into its association with vaccine acceptability enhances our understanding of possible intervention strategies and policy reforms that can promote vaccine uptake and strengthen the resilience of the public health system.

## 2. METHODS

### 2.1 Data and sample

This study recruited a cohort of respondents to complete an initial structured phone interview (N=1,371), with follow-up bi-monthly panel surveys for six months (three rounds in total). Interviews lasted between 20 and 30 minutes, collecting information on COVID-19 prevention behaviors and barriers, knowledge of COVID-19 symptoms, care-seeking, vaccine acceptability, and demographic data. Data collection was through a structured questionnaire using CATI (Computer Assisted Telephone Interview). The cohort included those with phone access who were able to complete the survey via telephone interviews. The cohort were recruited from a sample of participants of a

COVID-19 Knowledge, Attitudes and Practices (KAP) baseline survey in Mozambique who consented to be contacted for future research (approximately 97% of baseline participants consented to be re-contacted). The panel sample was stratified by gender and province of residence to achieve approximately equal numbers of male and female respondents, and even geographical coverage across all provinces of the country.

The target sample size was estimated to result in 390 participants per region by the final wave, assuming a 10% loss to follow-up between the first and second wave, and then a 5% loss thereafter. Attrition from the cohort occurred if the participant was not contactable, unavailable for interview, or if they did not re-consent to participate in a given round of data collection. Sample size and other characteristics for the three waves of data may be found in the supplemental materials.

### 2.2 Key variables of interest

The questionnaire asked three questions about vaccine acceptability, constituting the outcome variables for the present analysis. The first related to the willingness to receive the vaccine: “If a vaccine to prevent COVID-19 were available today, would you get it?” with response options: “definitely get it”, “probably get it”, “probably not”, and “I do not know”. We created a binary variable: 1 = definitely get it and 0 = other responses. Perceived efficacy was captured with the question: “what is your level of concern that a COVID-19 vaccine does not prevent the disease?”, with response options: “very worried”, “worried”, “not worried”, and “I believe the vaccine would prevent the disease”. We created a binary variable: 0 = (very) worried and 1= not worried or believing in the vaccine. The third question asked: “how worried are you that a COVID-19 vaccine is not safe?”, with response options: “very worried”, “worried, “not worried”, and “I believe the vaccine is safe”. We created a binary variable: 0 = (very) worried and 1 = not worried or believing in the vaccine’s safety. We created a summary measure for the acceptability of the COVID-19 vaccine by adding together these three binary variables.

The key independent variable of interest is institutional trust. The questionnaire included a six-item institutional trust battery, based on the WHO’s COVID-19 Global Risk Communication and Community Engagement Strategy guidance (11). Respondents were asked how much confidence they had with elected entities, government, local government (municipality), doctors, journalists, and business leaders to act in the public interest, with Likert-type response options: “no confidence”, “little confidence”, “some confidence”, and “a lot of confidence”, coded from 1 to 4. The Cronbach alpha of the six items is 0.77, indicating good internal consistency.

### 2.3 Control variables

We investigated a range of control variables, based on existing literature, that might confound the relationship between vaccine acceptability and institutional trust. Demographic factors included age, gender, and marital status. Age is a continuous variable. Marital status is a dichotomized variable: 0 = single/other (including never married, widowed, separated, divorced) and 1= currently married. Socioeconomic factors included education, employment, and the national wealth quintile (estimated using the Equity Tool) (12). We used a three-category education variable (primary or below, secondary, tertiary) while employment was dichotomized into 0 = unemployed and 1 = employed. A binary rural-urban residence variable was also analyzed.

### 2.4 Statistical analysis

We first conducted confirmatory factor analysis (CFA) to construct a latent score of institutional trust. The model specification was guided by model fit. We conducted the χ^2^ test, calculated the coefficient of determination (R^2^), and evaluated four fit indices including Root Mean Square Error of Approximation (RMSEA), Bentler Comparative Fit Index (CFI), Tucker-Lewis Index (TLI), and Standardized Root Mean Square Residual (SRMR). We calculated the predicted institutional trust score from the model with the best fit and normalized it to a scale ranging from 0 (no trust) to 100 (complete trust). The predicted score was included as an independent variable to predict vaccine acceptability.

The vaccine acceptability questions were asked in the three follow-up waves of the survey (namely waves 1-3). However, institutional trust questions were asked in the baseline, wave one and wave three of the survey. To make full use of the information available, using CFA we calculated the institutional trust score of the baseline, wave one and wave three surveys, and treated the mean score of the other three waves as a proxy for the trust score for wave two.

For the three binary acceptability variables, we built multilevel logistic regression models with random intercept (i.e., panel data random effects models) which account for individual-level heterogeneity. The overall acceptability variable is a count variable, so we built multilevel Poisson regression models with random intercept. We added time dummy variables in the regression models and calculated the predicted acceptability across different waves to map out the trajectories of vaccine acceptability. We conducted likelihood ratio tests and calculated the intra-class correlation to evaluate the usefulness of a multilevel model.

### 2.5 Ethical Approval

This study was determined to be human subjects research and received IRB approval from the Mozambique Comité Nacional de Bioética para Saúde (CNBS) (reference: 439/CNBS/20). All participants gave their informed consent to participate in the study.

## 3. RESULTS

The sample size in wave 1 was 1,327 falling to 1,185 by wave 3. The average age of the sample was 32 years old. Men accounted for 61% of the sample, over half (54%) of the sample were married. The mean value of the equivalized income score was 1.3. A majority of the participants of the survey received secondary (66%) or tertiary education (29%). Around 30% were unemployed, and 81% were living in urban areas (Table 1).

**Table 1.** Sample characteristics (N=3,809)

|  | Wave 1 | Wave 2 | Wave 3 | Waves 1-3 |
| --- | --- | --- | --- | --- |
|  | Proportions or means |  |  |  |
| Definitely Take vaccine |  |  |  |  |
| No | 63% | 56% | 71% | 63% |
| Yes | 37% | 44% | 29% | 37% |
| Vaccine efficacy |  |  |  |  |
| Worried | 67% | 67% | 72% | 69% |
| Not worried | 33% | 33% | 28% | 31% |
| Vaccine safety |  |  |  |  |
| Worried | 73% | 70% | 77% | 73% |
| Not worried | 27% | 30% | 23% | 27% |
| Vaccine acceptability score | 1.0 | 1.1 | 0.8 | 0.9 |
| Institutional trust score | 72.5 | 73.5 | 75.4 | 73.7 |
| Age | 32.3 | 32.3 | 32.5 | 32.3 |
| Gender |  |  |  |  |
| Female | 41% | 39% | 38% | 39% |
| Male | 59% | 61% | 62% | 61% |
| Marital status |  |  |  |  |
| Single | 47% | 48% | 43% | 46% |
| Married | 53% | 52% | 57% | 54% |
| Income score | 1.3 | 1.3 | 1.3 | 1.3 |
| Education |  |  |  |  |
| Primary education | 6% | 6% | 6% | 6% |
| Secondary education | 66% | 66% | 65% | 66% |
| Tertiary education | 28% | 28% | 29% | 29% |
| Employment |  |  |  |  |
| Unemployed | 31% | 25% | 30% | 29% |
| Employed | 69% | 75% | 70% | 71% |
| Rural/urban residence |  |  |  |  |
| Rural areas | 19% | 19% | 20% | 19% |
| Urban areas | 81% | 81% | 80% | 81% |
| N | 1,327 | 1,253 | 1,185 | 3,809 |

The results of the confirmatory factor analysis show that all six items in the institutional trust battery loaded significantly (*p*<0.001) and positively on a single-dimensional construct (Figure 1). Factor loadings for confidence in the government, local governments, and elected entities were higher than those for doctors, journalists, and business leaders. An increase of one standard deviation in institutional trust was associated with an increase of 0.88 standard deviations in confidence in the government, 0.72 standard deviations in confidence in local governments, and 0.69 standard deviations in confidence in elected entities. There are strong correlations between the residuals in relation to trust in governments, doctors, journalists, and business leaders. The post-estimation diagnostic tests show that the model has an excellent fit to the data. The χ^2^ test is not statistically significant, indicating little difference between the model-implied and sample covariance matrices. Both the RMSEA and SRMR are below the threshold of 0.05, and both the CFI and TLI are above the threshold of 0.95. The R^2^ statistic suggests that the six items explain 87% of the total variance of the latent construct.

**Figure 1.**
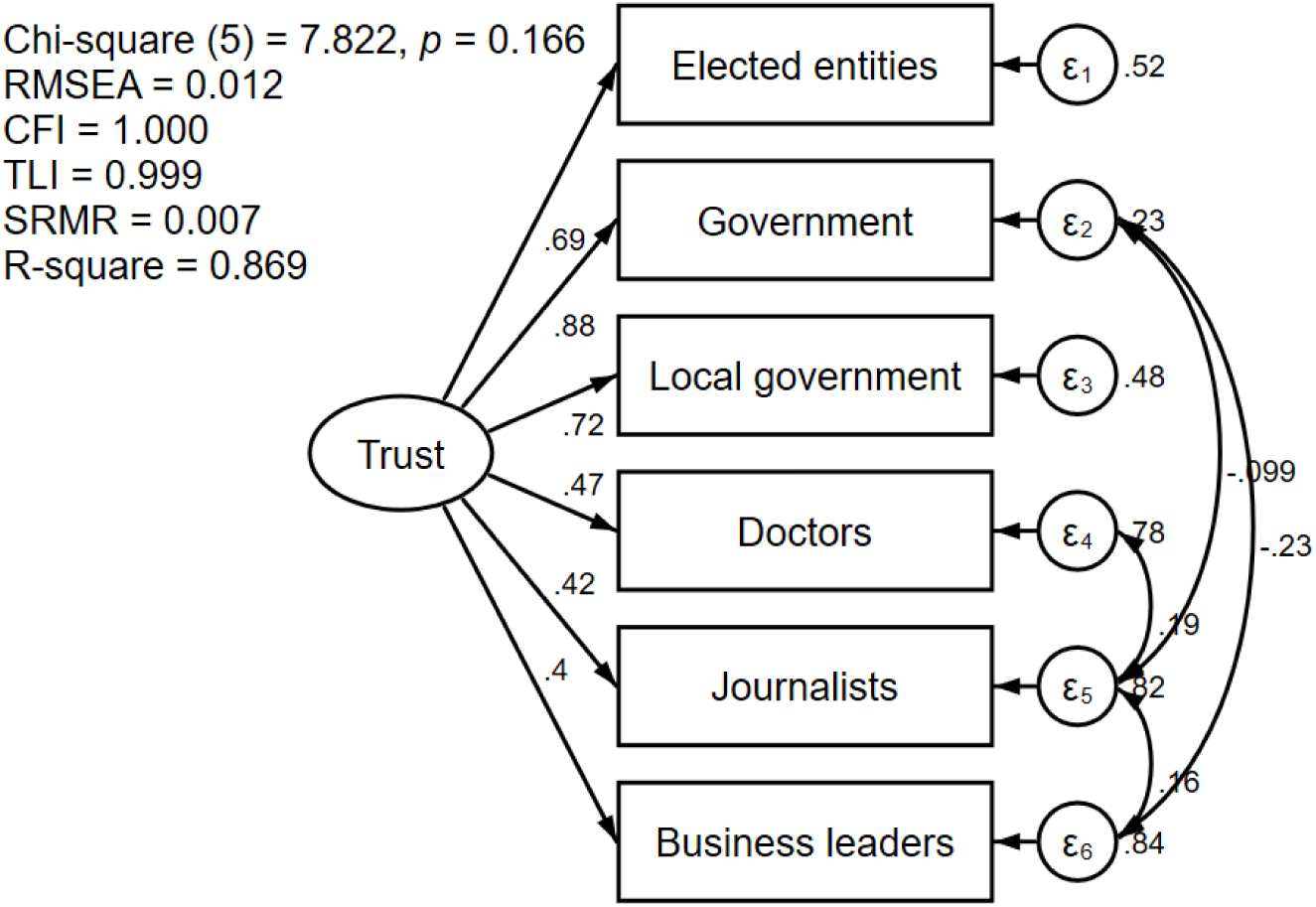
Factor analysis of institutional trust Notes: Standardized factor loadings are reported; RMSEA: Root Mean Square Error of Approximation; CFI: Comparative Fit Index; TLI: Tucker-Lewis Index; SRMR: Standardized Root Mean Square Residual; R-square: coefficient of determination.

There was a small increase in the mean institutional trust score between waves 1 and 3 (from 72.5 to 75.4, respectively) (Table 1). The percentage of respondents reporting they would definitely take the vaccine showed considerable variability over time, from 37% to 44% to 29% between waves 1, 2 and 3. There was an increase in concern about vaccine efficacy between waves 2 and 3: 33% reported that they were not worried about the efficacy of the vaccine in waves 1 and 2, decreasing to 28% in wave 3. Over-time variability was also seen in the percentage reporting they were not worried about the vaccine’s safety between waves 1, 2 and 3 (27%, 30%, and 23%, respectively). Similarly, the mean vaccine acceptability score increased from 1.0 in wave 1 to 1.1 in wave 2 before decreasing to 0.8 in wave 3.

The three dichotomized measures of vaccine acceptability were included as dependent variables in three separate regression models (Table 2). The institutional trust score is a statistically significant variable in all three models. A one-unit increase in the trust score was associated with an increase in the odds of: willingness to take the vaccine (17%; *p*<0.001); perceived vaccine efficacy (7%; *p*<0.01); and perceived vaccine safety (10%; *p*<0.001), controlling for other variables. Respondents reported a significantly higher level of willingness to take the vaccine in wave 2 than in wave 1, and a significantly lower level of willingness to take the vaccine, perceived vaccine efficacy, and perceived vaccine safety in wave 3. Older age or being male was significantly associated with increased willingness to take the vaccine, and perceived vaccine safety. Perceived vaccine efficacy did not differ significantly by gender, age or marital status. People in urban areas had a lower level of willingness to take the vaccine. Both the likelihood ratio test and the intra-class correlation coefficients suggest that vaccine acceptability was correlated within participants over time, indicating that a multilevel modelling approach was appropriate.

**Table 2.**
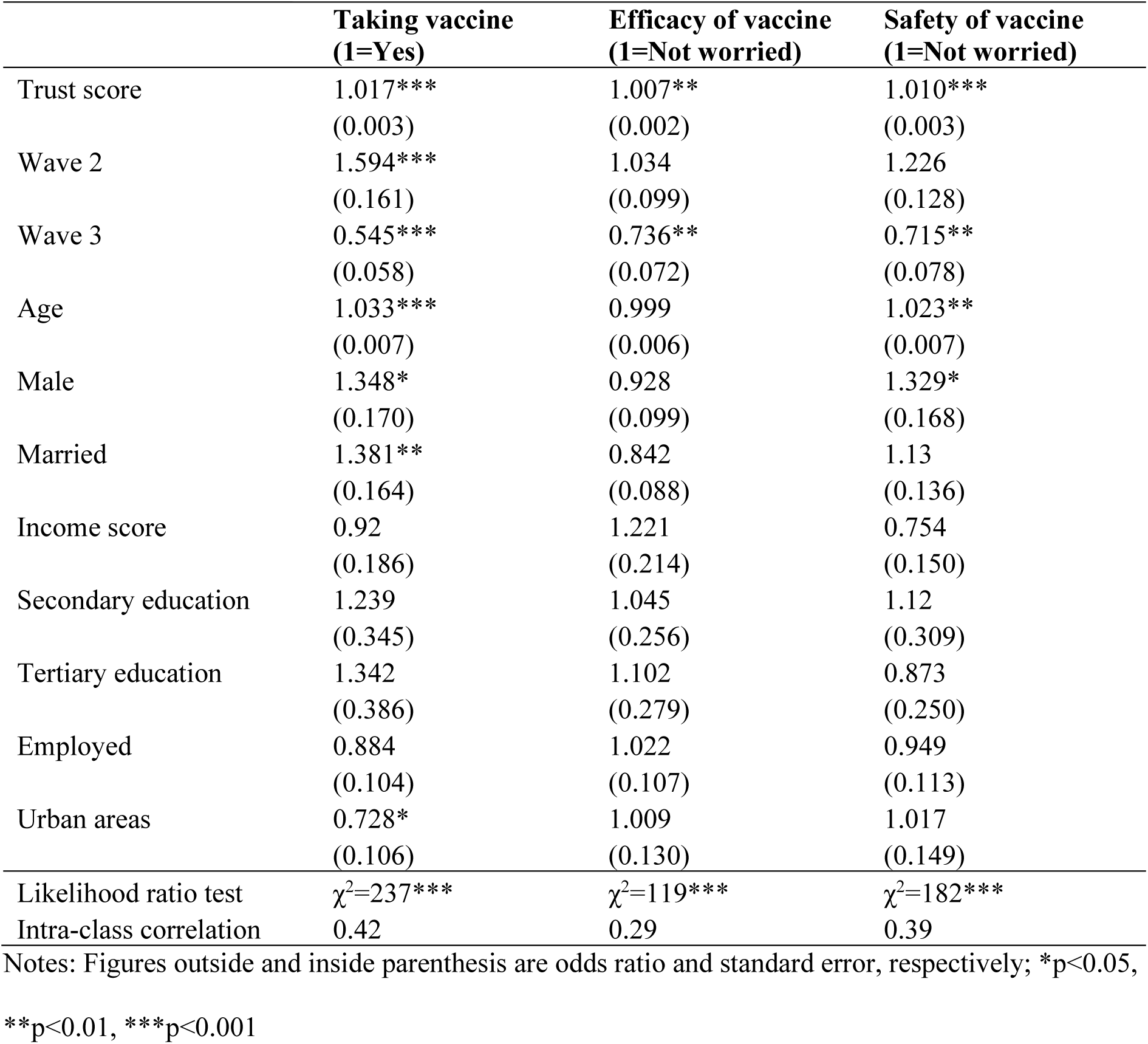
Multilevel binary logistic regression analysis of vaccine acceptability (N=3,741)

|  | **Taking vaccine**<br>**(1=Yes)** | **Efficacy of vaccine**<br>**(1=Not worried)** | **Safety of vaccine**<br>**(1=Not worried)** |
| --- | --- | --- | --- |
| Trust score | 1.017***<br>(0.003) | 1.007**<br>(0.002) | 1.010***<br>(0.003) |
| Wave 2 | 1.594***<br>(0.161) | 1.034<br>(0.099) | 1.226<br>(0.128) |
| Wave 3 | 0.545***<br>(0.058) | 0.736**<br>(0.072) | 0.715**<br>(0.078) |
| Age | 1.033***<br>(0.007) | 0.999<br>(0.006) | 1.023**<br>(0.007) |
| Male | 1.348*<br>(0.170) | 0.928<br>(0.099) | 1.329*<br>(0.168) |
| Married | 1.381**<br>(0.164) | 0.842<br>(0.088) | 1.13<br>(0.136) |
| Income score | 0.92<br>(0.186) | 1.221<br>(0.214) | 0.754<br>(0.150) |
| Secondary education | 1.239<br>(0.345) | 1.045<br>(0.256) | 1.12<br>(0.309) |
| Tertiary education | 1.342<br>(0.386) | 1.102<br>(0.279) | 0.873<br>(0.250) |
| Employed | 0.884<br>(0.104) | 1.022<br>(0.107) | 0.949<br>(0.113) |
| Urban areas | 0.728*<br>(0.106) | 1.009<br>(0.130) | 1.017<br>(0.149) |
| Likelihood ratio test | $\chi^2=237***$ | $\chi^2=119***$ | $\chi^2=182***$ |
| Intra-class correlation | 0.42 | 0.29 | 0.39 |

Notes: Figures outside and inside parenthesis are odds ratio and standard error, respectively; \*p<0.05,
\*\*p<0.01, \*\*\*p<0.001

The results from the multilevel Poisson regression model (Table 3) show that model 1 is largely consistent with the findings in Table 2. People with a higher institutional trust score, older people, and men had significantly greater vaccine acceptability. Acceptability increased significantly in wave 2 and then decreased significantly in wave 3. Since age and time dummies are consistently strong predictors of vaccine acceptability, we further investigated the interaction effects between institutional trust and these two predictors, to detect any non-linear relationships between institutional trust and vaccine acceptability and to reveal the heterogeneity in those relationships for people in different groups.

**Table 3.** Multilevel Poisson regression analysis of vaccine acceptability (N=3,741)

|  | **Model 1** | **Model 2** | **Model 3** |
| --- | --- | --- | --- |
| Trust score | 1.006***<br>(0.001) | 1.016***<br>(0.003) | 1.013***<br>(0.004) |
| Wave 2 | 1.115**<br>(0.044) | 1.115**<br>(0.044) | 0.955<br>(0.147) |
| Wave 3 | 0.804***<br>(0.035) | 0.805***<br>(0.035) | 0.491***<br>(0.080) |
| Age | 1.008***<br>(0.002) | 1.033***<br>(0.009) | 1.033***<br>(0.009) |
| Trust × Age |  | 0.9997**<br>(0.000) | 0.9997**<br>(0.000) |
| Trust × Wave 2 |  |  | 1.002<br>(0.002) |
| Trust × Wave 3 |  |  | 1.006**<br>(0.002) |
| Male | 1.088*<br>(0.047) | 1.084<br>(0.046) | 1.084<br>(0.046) |
| Married | 1.045<br>(0.044) | 1.033<br>(0.044) | 1.037<br>(0.044) |
| Income score | 0.979<br>(0.066) | 0.977<br>(0.066) | 0.976<br>(0.066) |
| Secondary education | 1.066<br>(0.100) | 1.036<br>(0.097) | 1.035<br>(0.097) |
| Tertiary education | 1.052<br>(0.102) | 1.021<br>(0.099) | 1.019<br>(0.099) |
| Employed | 0.971<br>(0.042) | 0.973<br>(0.042) | 0.973<br>(0.042) |
| Urban areas | 0.943<br>(0.047) | 0.944<br>(0.047) | 0.943<br>(0.047) |

Notes: Figures outside and inside parenthesis are incidence rate ratio and standard error, respectively;
\*p<0.05, \*\*p<0.01, \*\*\*p<0.001

Age had a significant moderation effect (model 2 and Figure 2a). The incidence rate ratio (IRR) of the interaction term indicates that the positive correlation between vaccine acceptability and institutional trust was weakened among people in older age groups. For people aged between 18 and 29 years, the predicted score of vaccine acceptability increases by 130%, from 0.47 to 1.08, as the institutional trust score rises from 0 to 100. This stands in contrast to the predicted acceptability score of people aged 40 and over which increases by 15%, from 1.01 to 1.16.

**Figure 2:**
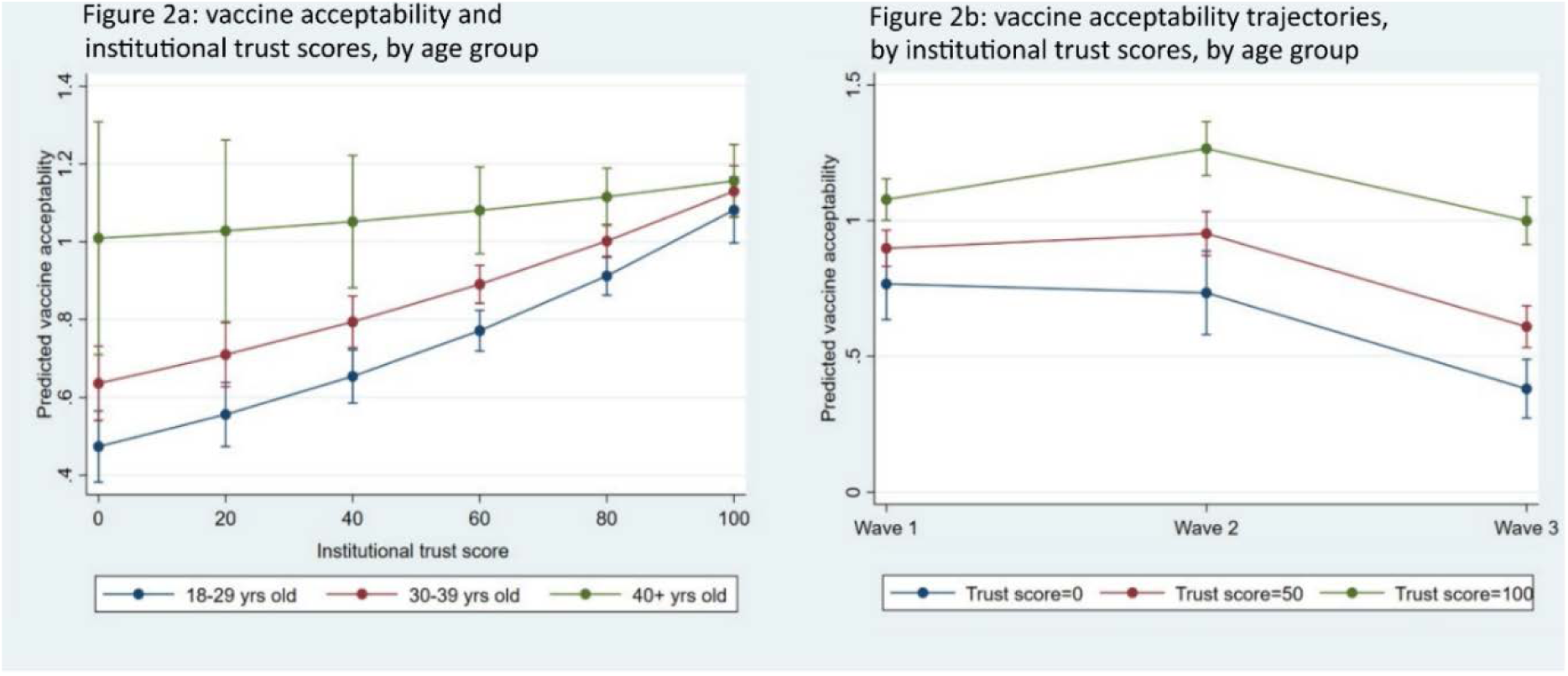
Predicted vaccine acceptability (95% confidence interval plotted on the mean) (N=3741)

Model 3 (Table 3) included an interaction term between institutional trust and time. Like in models 1 and 2, the IRR for wave 3 is significantly below 1, indicating a marked decrease in vaccine acceptability at that time point, while the IRR for the interaction between trust and wave 3 is significant and above 1. This means that the decrease in vaccine acceptability from wave 1 to 3 was attenuated among people with high institutional trust (Figure 2b). For people with a trust score of 50, the predicted acceptability score reduced from 0.90 in wave 1 to 0.61 in wave 3. For people with a trust score of 100, the predicted acceptability score increased from 1.01 in wave 1 to 1.26 in wave 2 before reducing to 1.00 in wave 3. The large decrease in vaccine acceptability between wave 1 and 3 was mainly driven by people with low institutional trust.

## 4. DISCUSSION

This study is among the first to examine the relationship between institutional trust and COVID-19 vaccination acceptability. We have several new and compelling findings. First, we found that vaccination acceptability was volatile over the study period of six months. Acceptability increased from September to November 2020, but decreased by the final wave in January-February 2021. One possible explanation is that negative news regarding vaccination safety and efficacy was released by major local and international media around the time of the final data collection period. Evidence from Japan, Portugal and the UK showed that vaccination acceptability was highly associated with the perceived safety and efficacy of the vaccinations (12–14). The main reasons for receiving vaccinations are for people to protect themselves and others from COVID-19. Therefore, when uncertainties around vaccination safety and efficacy arise, it is important to engage the public with transparency and accountability, to prevent uncertainties from transforming into negative perceptions.

Based on a score we constructed in the survey, we found that higher institutional trust was significantly associated with higher vaccine acceptability. This finding is consistent with the broader literature on social and political determinants of acceptability for vaccination. It is shown, for example, that trust in federal institutions (15), and trust in the government’s technical and organizational skill to handle a contagious outbreak along with confidence in medical institutions play a salient role in predicting willingness to be vaccinated against influenza in the US (16). Similar results have been seen in the UK, linking positive attitudes towards vaccination with a high level of trust in the National Health Service and the government (17). In addition to contributing to this literature for the Global South, our study additionally uses longitudinal data, which shows that people with a high level of trust are less susceptible to a decline in vaccine acceptability over time. This finding seems to suggest that the public’s trust in institutions can contribute to more resilience in the public health system in terms of vaccine uptake and health service utilization in the context of a global pandemic.

Mozambique has invested considerable efforts toward reliable sources of information, aiming to overcome misinformation and bias against vaccination programs. This approach has involved key figures in the ministry of health, public health and research institutions addressing the public on a daily basis. Scientific virtual conferences on the COVID-19 pandemic have included vaccines and were implemented with renowned and trusted persons taking a prominent role. Scientific conferences and media appearances were broadcast both on radio and television. These informational and SBCC efforts to build trust in the general population seem well-placed in light of our findings linking greater institutional trust with increased vaccine acceptability. During an active pandemic, these examples in Mozambique represent strong cross-institutional collaboration across government, media, academics, and public health practitioners. We recommend that the Ministry of Health and their partners continue to cultivate such collaboration before, during, and after emerging pandemic threats to ensure robust health system responses that routinely nurture or instill trust, which we found to be highly variable.

We found that some demographic factors predicted vaccine acceptability. This is consistent with another study based on an online health worker survey on vaccination acceptability in Mozambique which found that vaccination acceptability was positively and significantly associated with age (9), although existing studies examining gender differences in vaccine acceptability have reported mixed results (9,13). Inequities in vaccination acceptability (and thus likely uptake) reflect the need for a greater exploration of its drivers and for interventions to address potential inequalities to allow for more equitable uptake of vaccinations in future. Targeted campaigns to promote vaccination acceptability among those groups with low levels of trust are needed.

Our results have to be interpreted in light of the study limitations. The data were collected remotely through telephone interviews. This means that our sample consisted solely of people with access to a cell phone, who are also likely to have greater access to health care. These people represent a subset of the target population, and the sample is likely to be biased towards urban, wealthier respondents. Our recruitment strategy achieved a high response rate, but our sample was not representative of the general population so any generalization should be made with caution.

## 5. CONCLUSION

Our study underscores the central role of health communication and trust-building in promoting vaccine acceptability to protect public health. Although personal characteristics such as gender, age, and marital status have an influence on vaccine uptake, the effects of broader structural factors, including trust in institutions, should not be ignored. Simply making vaccines available is not sufficient to ensure access and uptake. Building a resilient health system requires proactive engagement with the public, building and maintaining a trusting relationship between healthcare users and institutions. There is considerable variability in societal attitudes towards vaccine programs, even over short periods of time. Finding ways to build and capitalize on trust in institutions, particularly among those less likely to vaccinate, could prove successful in increasing vaccine acceptability in many contexts, and help underpin the resilience of the health system.

## Data Availability

The data consist of deidentified participant data and are available upon reasonable request from the corresponding author subject to PSIs data use policy

## ACKNOWLEDGEMENTS

We would like to acknowledge the generosity of the study participants in sharing their time and information with us, often repeatedly over many months. We would also like to thank the field staff working tirelessly during a pandemic to call participants and gather these data.

## PATIENT AND PUBLIC INVOLVEMENT

This study did not collect data from patients specifically. Patients and the public were not involved in the study. However, de-identified data is available to the public on request.

## DATA AVAILABILITY STATEMENT

The data consist of deidentified participant data, and are available upon reasonable request from the corresponding author, subject to PSI’s data use policy.

## SUPPLEMENTAL MATERIALS

**Supplemental table 1:**
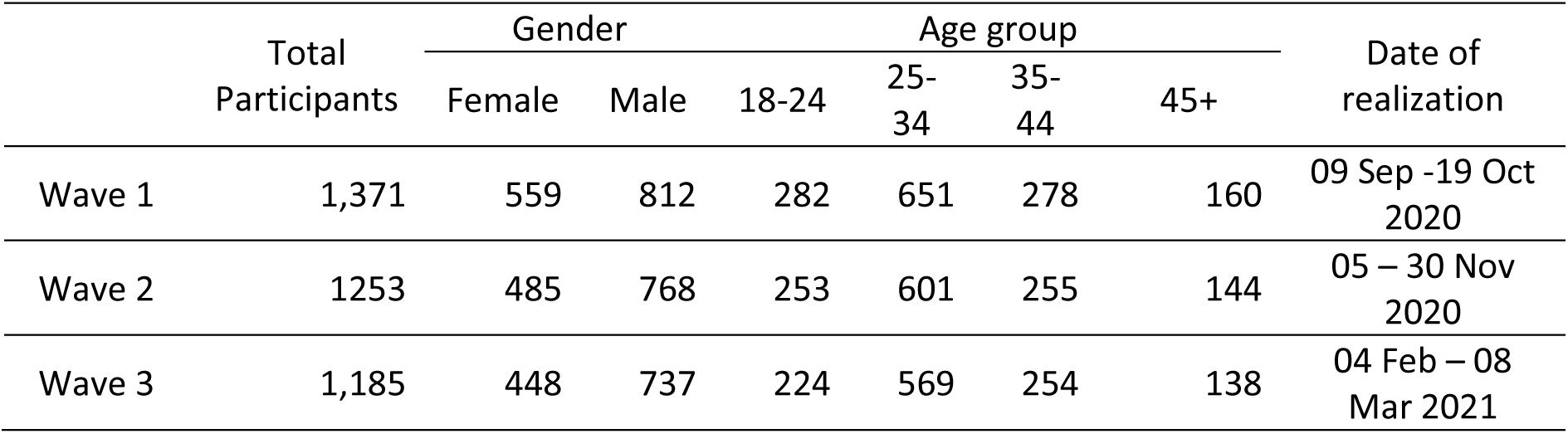
Data collection dates, sample sizes and basic characteristics.

|  | Total<br>Participants | Gender |  | Age group |  |  |  | Date of<br>realization |
| --- | --- | --- | --- | --- | --- | --- | --- | --- |
|  |  | Female | Male | 18-24 | 25-34 | 35-44 | 45+ |  |
| Wave 1 | 1,371 | 559 | 812 | 282 | 651 | 278 | 160 | 09 Sep -19 Oct<br>2020 |
| Wave 2 | 1253 | 485 | 768 | 253 | 601 | 255 | 144 | 05 – 30 Nov<br>2020 |
| Wave 3 | 1,185 | 448 | 737 | 224 | 569 | 254 | 138 | 04 Feb – 08<br>Mar 2021 |

## REFERENCES

1. Bok K, Sitar S, Graham BS, Mascola JR. Accelerated COVID-19 vaccine development: milestones, lessons, and prospects. Immunity. 2021 Aug 10;54(8):1636–51.

2. Mellet J, Pepper MS. A COVID-19 Vaccine: Big Strides Come with Big Challenges. Vaccines (Basel). 2021 Jan 11;9(1):39.

3. Machingaidze S, Wiysonge CS. Understanding COVID-19 vaccine hesitancy. Nat Med. 2021 Aug;27(8):1338–9.

4. World Health Organization. 10 global health issues to track in 2021 [Internet]. [cited 2022 Jan 27]. Available from: https://www.who.int/news-room/spotlight/10-global-health-issues-to-track-in-2021

5. Wilson SL, Wiysonge C. Social media and vaccine hesitancy. BMJ Global Health. 2020 Oct 1;5(10):e004206.

6. Raude J, Fressard L, Gautier A, Pulcini C, Peretti-Watel P, Verger P. Opening the ‘Vaccine Hesitancy’ black box: how trust in institutions affects French GPs’ vaccination practices. Expert Rev Vaccines. 2016 Jul;15(7):937–48.

7. Palamenghi L, Barello S, Boccia S, Graffigna G. Mistrust in biomedical research and vaccine hesitancy: the forefront challenge in the battle against COVID-19 in Italy. Eur J Epidemiol. 2020 Aug;35(8):785–8.

8. Robertson E, Reeve KS, Niedzwiedz CL, Moore J, Blake M, Green M, et al. Predictors of COVID-19 vaccine hesitancy in the UK household longitudinal study - ScienceDirect. Brain, Behavior, and Immunity. 94(1):41–50.

9. Dula J, Mulhanga A, Nhanombe A, Cumbi L, Júnior A, Gwatsvaira J, et al. COVID-19 Vaccine Acceptability and Its Determinants in Mozambique: An Online Survey. Vaccines. 2021 Aug;9(8):828.

10. COVID-19 Global Risk Communication and Community Engagement Strategy – interim guidance [Internet]. [cited 2021 Dec 3]. Available from: https://www.who.int/publications-detail-redirect/covid-19-global-risk-communication-and-community-engagement-strategy

11. Equity Tool Metrics. Equity Tool for Mozambique [Internet]. Equity Tool. [cited 2021 Dec 3]. Available from: https://www.equitytool.org/mozambique/

12. Bell S, Clarke R, Mounier-Jack S, Walker JL, Paterson P. Parents’ and guardians’ views on the acceptability of a future COVID-19 vaccine: A multi-methods study in England. Vaccine. 2020 Nov 17;38(49):7789–98.

13. Soares P, Rocha JV, Moniz M, Gama A, Laires PA, Pedro AR, et al. Factors Associated with COVID-19 Vaccine Hesitancy. Vaccines. 2021 Mar;9(3):300.

14. Machida M, Nakamura I, Kojima T, Saito R, Nakaya T, Hanibuchi T, et al. Acceptance of a COVID-19 Vaccine in Japan during the COVID-19 Pandemic. Vaccines. 2021 Mar;9(3):210.

15. Jamison AM, Quinn SC, Freimuth VS. “You don’t trust a government vaccine”: Narratives of institutional trust and influenza vaccination among African American and white adults. Social Science & Medicine. 2019 Jan 1;221:87–94.

16. Mesch GS, Schwirian KP. Social and political determinants of vaccine hesitancy: Lessons learned from the H1N1 pandemic of 2009-2010. American Journal of Infection Control. 2015 Nov 1;43(11):1161–5.

17. Sethi S, Kumar A, Mandal A, Shaikh M, Hall CA, Kirk JMW, et al. The UPTAKE study: a cross-sectional survey examining the insights and beliefs of the UK population on COVID-19 vaccine uptake and hesitancy. BMJ Open. 2021 Jun 1;11(6):e048856.

